# Changes in isolation guidelines for CPE patients results in only a mild reduction in required hospital beds

**DOI:** 10.1101/2024.07.04.24309973

**Authors:** Michael J Lydeamore, David Wu, Tjibbe Donker, Claire Gorrie, Charlie K Higgs, Marion Easton, Daneeta Hennessy, Nicholas Geard, Benjamin P Howden, Ben S Cooper, Andrew Wilson, Anton Y Peleg, Andrew J Stewardson

**Affiliations:** Department of Econometrics and Business Statistics, Monash University, Clayton VIC Australia; Victorian Department of Health, Government of Victoria, Melbourne, VIC Australia; Institute for Infection Prevention and Hospital Epidemiology, University Medical Center Freiburg, Germany; Department of Microbiology & Immunology, University of Melbourne, at the Peter Doherty Institute for Infection and Immunity, Melbourne, VIC Australia; Microbiological Diagnostic Unit Public Health Laboratory, Department of Microbiology & Immunology, University of Melbourne, at the Peter Doherty Institute for Infection and Immunity, Melbourne, VIC Australia; School of Computing and Information Systems, Faculty of Engineering and Information Technology, University of Melbourne, Melbourne, VIC Australia; Centre for Tropical Medicine and Global Health, Nuffield Department of Medicine, University of Oxford, United Kingdom; Department of Infectious Diseases, The Alfred and School of Translational Medicine, Monash University, Melbourne, VIC Australia; Infection Program, Monash Biomedicine Discovery Institute, Department of Microbiology, Monash University, Melbourne, VIC Australia; Centre to Impact AMR, Monash University, Melbourne, VIC Australia

## Abstract

Carbapenemase-producing Enterobacterales (CPE) are an emerging public health concern globally as they are resistant to a broad spectrum of antibiotics. Colonisation with CPE typically requires patients to be managed under ‘contact precautions’, which creates additional physical bed demands in healthcare facilities. This study examined the potential impact of revised isolation guidelines introduced in late 2023 in Victoria, Australia, that relaxed the requirement for indefinite isolation of CPE-colonised patients in contact precautions, based on admission of CPE-diagnosed cases prior to the guideline change. Our analysis showed that while the changes result in modest savings in the need for dedicated isolation rooms, they could reduce the duration of time individual patients spend in isolation by up to three weeks. However, ongoing investments to expand isolation capacity would still be required to accommodate the rising incidence of CPE.

## 1. Introduction

Carbapenemase-producing *Enterobacterales* (CPE) present a serious health threat due to the high morbidity and mortality associated with infection^1^. In Victoria, Australia, CPE infections have been steadily increasing in recent years despite a centralised ‘search and contain’ policy, with acquisition primarily associated with international travel and healthcare exposure^2^.

One of the practical challenges associated with rising CPE cases is the recommendation for colonised patients to be managed under ‘contact precautions’ when admitted to hospital. Specifically, the requirement for patients to be in a single room with ensuite (or dedicated bathroom) means that as the number of cases increases, so too does the number of single-bed rooms required for these patients to receive care. This is compounded by simultaneous increases in other conditions requiring a single room. As well as challenges related to the healthcare system, the requirement for contact precautions also places a significant burden on patients who can be isolated for long periods of time.

When first released in December 2015, the Victorian Guideline on Carbapenemase Producing Enterobacteriaceae for Health Services stated that patients colonised with CPE should be managed in contact precautions, including being isolated to a single room, regardless of subsequent screening results. This guidance persisted until a recent change whereby individuals can be risk assessed for release from contact precautions if 12 months have passed since their last positive CPE result^3^. Similar guidelines have been adopted internationally^4^.

We used a statewide linkage dataset to investigate the potential impact of this policy change on the demand for single rooms in Victorian hospitals. We compare the number of single rooms under the previous indefinite isolation policy to a best-case scenario under the new clearance criteria.

## 2. Methods

### Data sources

We used a line-listed, linked patient-level hospital admission dataset from the Evaluation and Control of Healthcare Infection Dissemination using Network Analysis (ECHIDNA) study. All patient admissions to Victorian public and private hospitals for the period 1 Jan 2011 to 31 Dec 2019 were sourced from the Victorian Admitted Episodes Dataset (VAED)^5^. CPE colonisation or infection status was obtained from the Public Health Event Surveillance System (PHESS) dataset, which records information on every notifiable condition in Victoria. CPE was made notifiable in Victoria since December 2019, and was recommended to be reported since 2016. Linkage between the PHESS and VAED datasets was performed by the Centre for Victorian Data Linkage, as part of the Victorian Linkage Map. All hospitals are included in this analysis, including day hospitals.

To determine the period of time a patient requires contact precautions while admitted under the revised guidance, we used the ‘calculated onset date’ of a notified patient (from PHESS), which is determined as follows: if the patient’s acquisition date was known, it was used as the calculated onset date; if the infection date was unavailable, the specimen collection date was used instead.

### Analysis

It is assumed contact precautions start on the calculated onset date for a patient. Under the previous policy (i.e. indefinite isolation), we assumed that the patient is in contact precautions for *all* future admissions. Under the best-case scenario for the current policy, we assumed that the patient is in contact precautions only for admissions that occur within 12 months of the calculated onset date. If a patient is in hospital on the 12-month date of diagnosis, we assumed they remain in contact precautions for that entire hospital stay. If a patient has more than one CPE notification, then the 12-month timer starts at the end of their latest calculated onset date.

### Ethics

Ethics permission was obtained from the Alfred Health ethics board, submission number 445/21.

## 3. Results

In the last full year of data (2019), under the indefinite isolation strategy, 18,798 patient-days required a single room for 3195 hospital admissions relating to 298 unique patients. Under the new strategy, 13,767 patient days would be required for 1517 hospital admissions relating to 177 unique patients. The change in clearance policy was therefore associated with a reduction in required single room bed-days of up to approximately 27% (Figure 1).

**Figure 1:**
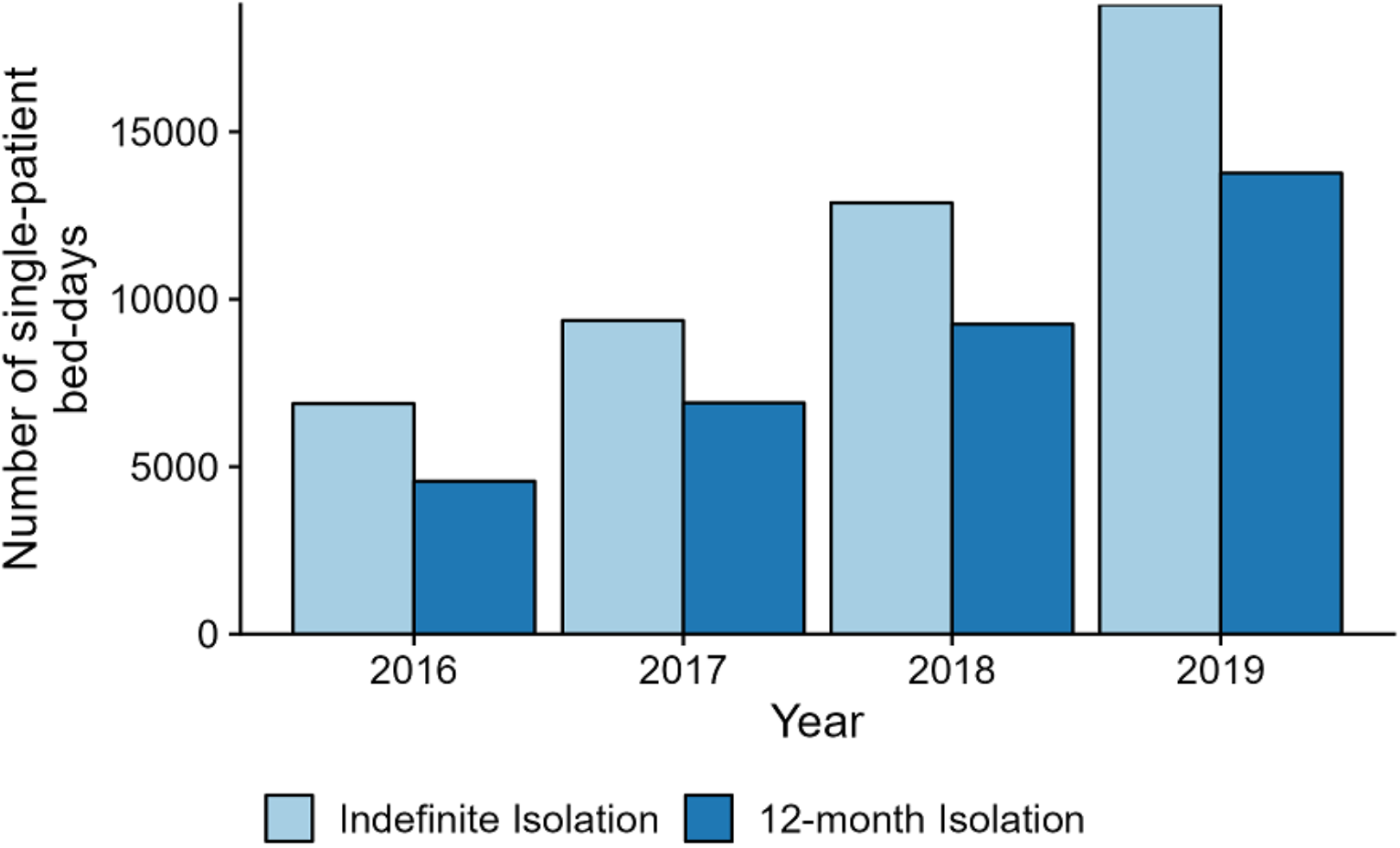
Number of single patient bed-days required in Victoria to accommodate the CPE colonised patients under two scenarios, by year. Scenarios are the prior Victorian recommendation that patients be isolated indefinitely (“always isolated”) and a best case scenario of the current guidance where patients may be risk assessed for “clearance” 12 months after their last CPE positive isolate.

Under the indefinite isolation strategy in the last full year of data (2019), patients were isolated for a median of 70 days (IQR 30-153). Under the new strategy, this would be reduced to a median of 46 days (IQR 13-99), meaning that each patient could potentially reduce their total time in isolation by more than three weeks.

Although this is a significant reduction in isolated bed-days for CPE patients, it can be seen in Figure 1 that, due to growth in CPE infections, the number of isolated bed-days under the newer strategy exceeds that of the previous year under the old (indefinite isolation) strategy. That is, the change in policy delays the increase in bed-days required by 12 months.

Grouping hospitals by peer group shows that the number of physical beds is not evenly spread across peer groups, with the majority of beds required in principal referral hospitals (Table 1). There is a modest reduction in each peer group under the new guidelines. Overall, this change in isolation criteria could result in 16 fewer isolated beds required.

**Table 1:** Maximum number of single rooms required to accommodate patients colonised by CPE, stratified by hospital peer group.

| Peer Group |  | Maximum required single rooms |  |
| --- | --- | --- | --- |
|  |  | *Indefinite isolation* | *No isolation after 12 months* |
| Public | Principal referral | 35 | 30 |
|  | Acute Group A | 10 | 7 |
|  | Acute Group B | 5 | 4 |
|  | Acute Group C | 2 | 2 |
| Private | Acute Group A | 6 | 5 |
|  | Acute Group B | 2 | 2 |
|  | Acute Group C | 1 | 1 |
| Mixed | Sub & non-acute | 13 | 7 |
| **Total** |  | **74** | **58** |

## 4. Discussion

This analysis shows that the change in guidelines requiring patients to no longer be in contact precautions 12 months after diagnosis in a best-case scenario could result in a systemwide saving of approximately 5000 bed-days in 12 months and require 16 fewer physical single room beds across the entire health system. From a patient perspective, the change in guidelines could also result in patients spending an average of 24 days less in isolation over their lifetime.

We found that the difference in bed-days over time between the two strategies continues to increase, as the cumulative number of patients ever diagnosed with CPE presenting to hospital also increases. This difference will not grow forever, as eventually patients diagnosed a long time ago with CPE will stop presenting to the healthcare system. However, in this setting, it seems as though the demand for single beds will continue to increase.

This work used several key assumptions that make these savings a best-case estimate. The first assumption is related to clearance criteria. For the updated scenario, we assumed that all CPE-colonised patients admitted more than 12 months from the date of CPE detection are cleared from the requirement for isolation. In practice, they would need three negative CPE screening samples, each collected at least 24 hours apart^3^. The second assumption is that patients diagnosed with CPE are always in contact precautions. In reality, without a centralised notification system, not all of these patients will be in contact precautions on each admission.

We evaluated the impact of a change in CPE clearance policy regarding demand for single rooms. Limited access to single rooms is a frequent challenge for health facilities given the multiple demands for these rooms, including various communicable diseases and other indications, such as end-of-life care. This policy change will likely result in a lesser demand of single rooms at least in the short term, but alone is unlikely to provide a sustained reduction in physical space requirements. We acknowledge, however, that an important implication of this policy change relates to patient experience, as the previous need for isolation precautions indefinitely comes with a significant burden on patients^6^.

## Data Availability

All data produced in the present study are available upon reasonable request to the authors, subject to relevant ethics guidelines and approvals.

## Acknowledgements

We would like to acknowledge the Victorian Department of Health as the source of the VAED and PHESS datasets for this study, and the Centre for Victorian Data Linkage (Victorian Department of Health) for the provision of the data linkage.

The opinions expressed are those of the authors and do not represent the views, nor the policy directions of the Victorian Department of Health.Competing interests

The authors declare no competing interests as part of this work.

